# Prevalence, clinical characteristics and treatment outcomes of HIV and SARS-CoV-2 co-infection: a systematic review and meta-analysis

**DOI:** 10.1101/2020.05.31.20118497

**Authors:** Joseph Baruch Baluku, Ronald Olum, Curthbert Agolor, Josephine Nakakande, Laura Russell, Felix Bongomin, Jane Nakawesi

## Abstract

**Objectives:** To determine the prevalence, clinical characteristics and outcomes of HIV and severe acute respiratory syndrome coronavirus 2 (SARS-CoV-2) co-infection.

**Methods:** We searched Medline, Embase, Cochrane and Web of Science databases and grey literature for studies reporting epidemiological and clinical data of patients with HIV and SARS-CoV-2 co-infection. Eligible studies were all observational or interventional studies and commentaries in English language that reported patient data on HIV/SARS-CoV-2 co-infection. We used random effect meta-analysis to determine the pooled prevalence and mortality.

**Results:** Of the 17 eligible studies, there were 3 retrospective cohorts, 1 survey, 5 case series, 7 case reports and 1 commentary that reported on a total of 146 HIV infected individuals. The pooled prevalence of HIV among individuals with SARS-CoV-2 infection was 1.0% (95% CI: 0.0 – 3.0, I^2^ = 79.3%, p = 0.01), whereas the prevalence of SARS-CoV-2 among HIV patients was 0.68% (95% CI: 0.34 – 1.34).There were 110 (83.8%) HIV/ SARS-CoV-2 co-infected males, and the age (range) of the co-infected was 30 – 60 years. A total of 129 (97.0%) were anti-retroviral therapy experienced, and 113 (85.6%) had a suppressed HIV viral load. The CD4 count (range) was 298 – 670 cells/mm^3^ (n = 107). The commonest symptoms were fever (73.5%, n = 75) and cough (57.8%, n = 59). Sixty-two (65.3%) patients had at least one other comorbid condition, of which hypertension (26.4%, n = 38) was the commonest. Chest radiological imaging abnormalities were found in 46 (54.1%) cases. Twenty-eight cases (56.0%) were reported as mild. Recovery occurred in 120 (88.9%) cases, and the pooled mortality was 9% (95% CI: 3.0 – 15.0, I^2^ = 25.6%, p = 0.24).

**Conclusion:** The prevalence of HIV/SARS-CoV-2 co-infection was low. The clinical characteristics and outcomes of HIV/SARS-CoV-2 co-infection are comparable to those reported among HIV negative SARS-CoV-2 cases.

## BACKGROUND

Over 5.5 million cases of coronavirus disease of 2019 (COVID-19) – caused by the severe acute respiratory syndrome coronavirus 2 (SARS-CoV-2) – have been reported globally with approximately 350,000 deaths as of 28^th^ May 2020 (1). Pre-existing conditions such as respiratory disease, hypertension and cardiovascular disease have been linked to severe forms of COVID – 19 (2). Epidemiological and clinical implications of HIV/SARS-CoV-2 co-infection are unknown for the 38 million people living with HIV (PLHIV) globally (3). SARS-CoV-2 infection has been reported to cause immune dysregulation that is characterised by low CD4 counts and elevated interleukin-6 (IL – 6), both of which can theoretically increase the risk of superimposed opportunistic infections and death among PLHIV (4). Moreover, PLHIV have a high prevalence of the pre-existing conditions that are associated with poor outcomes of COVID – 19 (5). There is need for synthesis of the currently available reports on HIV/SARS-CoV-2 co-infection to guide HIV care programs and clinical teams in the management of PLHIV, especially in resource limited settings where 70% of PLHIV are found.

## OBJECTIVES

The aim of this systematic review and meta-analysis was to determine the prevalence, clinical characteristics and outcomes of HIV/SARS-CoV-2 co-infection.

## MATERIALS AND METHODS

### Data sources, study eligibility and selection

This study was performed according to the Preferred Reporting Items for Systematic Reviews and Meta-Analyses (PRISMA) guidelines (6). We searched for clinical studies from Medline, Embase, Cochrane and Web of Science databases published in English from inception to May 22 2020. Duplicates were automatically removed using HDAS (https://hdas.nice.org.uk/). The search strategy is shown in appendix 1. There were no unique results on Cochrane or Web of Science databases. We also manually searched for additional sources from reference lists of eligible publications and pre-print repositories. Studies were reviewed by title and abstract by LR and JBB. Eligible studies were all observational or interventional studies and commentaries that reported data on patients with HIV and SARS-CoV-2 co-infection. For the meta-analysis of mortality, studies that reported data on less than 4 subjects were excluded. The study flow diagram is shown in figure 1.

**Figure 1:**
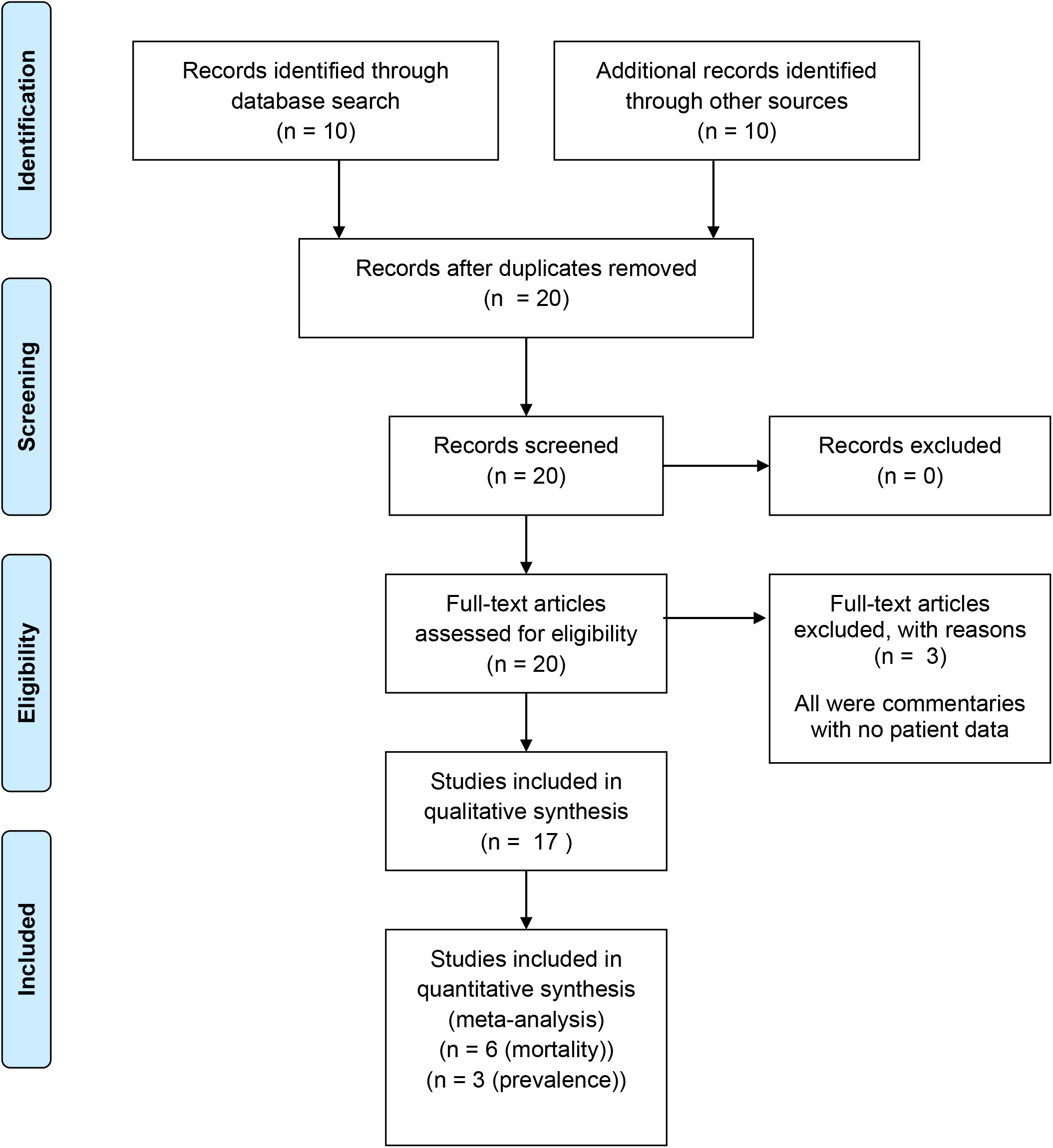
Study flow diagram

### Data extraction, study outcomes and analysis

From eligible studies, JBB and OR extracted data on study design, year of publication, country of origin and number of participants. We further extracted aggregated and/or individual patient data on prevalence of HIV among SARS-CoV-2 cases, prevalence of SARS-CoV-2 among PLHIV, demographic characteristics of HIV/SARS-CoV-2 co-infected patients, presenting complaints, other comorbid conditions, radiological chest imaging findings, disease severity, drugs used in management and treatment outcomes. Disagreements were resolved by a third reviewer (FB). The study outcomes were the prevalence of HIV/SARS-CoV-2 co-infection, the proportions of clinical characteristics and treatment outcomes of HIV/SARS-CoV-2 co-infection. We used random effect meta-analysis to determine pooled prevalence and mortality. Analysis was performed using STATA 16.0. Heterogeneity across studies was assessed using Q statistics and significance was set at p< 0.05 at the 95% confidence interval (CI).

## RESULTS AND DISCUSSION

### Study characteristics

As shown in table 1, 17 studies were included in the systematic review, of which 3 (7–9) and 6 (7,8,10–13) were included in the prevalence and mortality meta-analysis respectively. All studies were published in the year 2020. There were 3 retrospective cohorts, 1 survey, 5 case series, 6 case reports and 1 commentary. Of all studies, only 2 case reports (14,15) were from sub-Saharan Africa where 70% of HIV patients are found (16). While this may limit the generalizability of the findings of this review, it raises an urgent need for reports on HIV/SARS-CoV-2 co-infection from this region. The total number of HIV/SARS-CoV-2 coinfected patients reported on was 146.

**Table 1:** Characteristics of studies.

| **Study** | **Design** | **Country** | **No. of patients** |
| --- | --- | --- | --- |
| **Gervasoni, *et al.* (10)<sup>†</sup>** | Retrospective cohort | Italy | 47 |
| **Härter, *et al.* (11)** | Multicentre case series | Germany | 33 |
| **Karmen-Tuohy, *et al.* (7)** | Retrospective matched cohort | USA | 21 |
| **Okoh, *et al.* (8)** | Retrospective cohort | USA | 13 |
| **Guo, *et al.* (13)** | Survey | China | 8 |
| **Blanco, *et al.* (9)** | Case series | Spain | 5 |
| **Benkovic, *et al.* (22)** | Case series | USA | 4 |
| **Aydin, *et al.* (12)** | Case series | Turkey | 4 |
| **Riva, *et al.* (23)** | Case series | Italy | 3 |
| **Wang, *et al.* (27)** | Case report | China | 1 |
| **Drain, *et al.* (21)** | Commentary | USA | 1 |
| **Chen *et al.* (24)** | Case report | China | 1 |
| **Baluku *et al.* (15)** | Case report | Uganda | 1 |
| **Parker *et al.* (14)** | Case report | South Africa | 1 |
| **Louisa *et al.* (25)** | Case report | Singapore | 1 |
| **Zhu *et al.* (28)** | Case report | China | 1 |
| **Zhao *et al.* (18)** | Case report | China | 1 |

<sup>†</sup>includes 19 patients with probable SARS-CoV-2 but were not tested

### Prevalence of HIV/SARS-CoV-2 co-infection

Three studies (7–9) reported the proportion of HIV infected individuals among a total of 3,438 SARS-CoV-2 cases. The pooled prevalence of HIV among SARS-CoV-2 was 1.0% (95% confidence interval (CI): 0.0 – 3.0, I^2^ = 79.3%, p = 0.01) as shown in the forest plot in figure 2. There was significant heterogeneity of the studies and the estimate should be interpreted cautiously. Guo, *et al*., (13) reported a prevalence of SARS-Cov-2 of 0.68% (95% CI: 0.34 – 1.34) among 1,174 HIV patients. Gervasoni *et al*., (10) also reported to have identified 47 SARS-CoV-2 infected individuals from a database of “nearly” 6,000 PLHIV, thus giving an approximate prevalence of 0.78% (95% CI: 0.59 – 1.04). The prevalence of SARS-CoV-2 among PLHIV is therefore comparable to the 0.6 – 0.8% SARS-CoV-2 prevalence reported in the general population (17).

**Figure 2:**
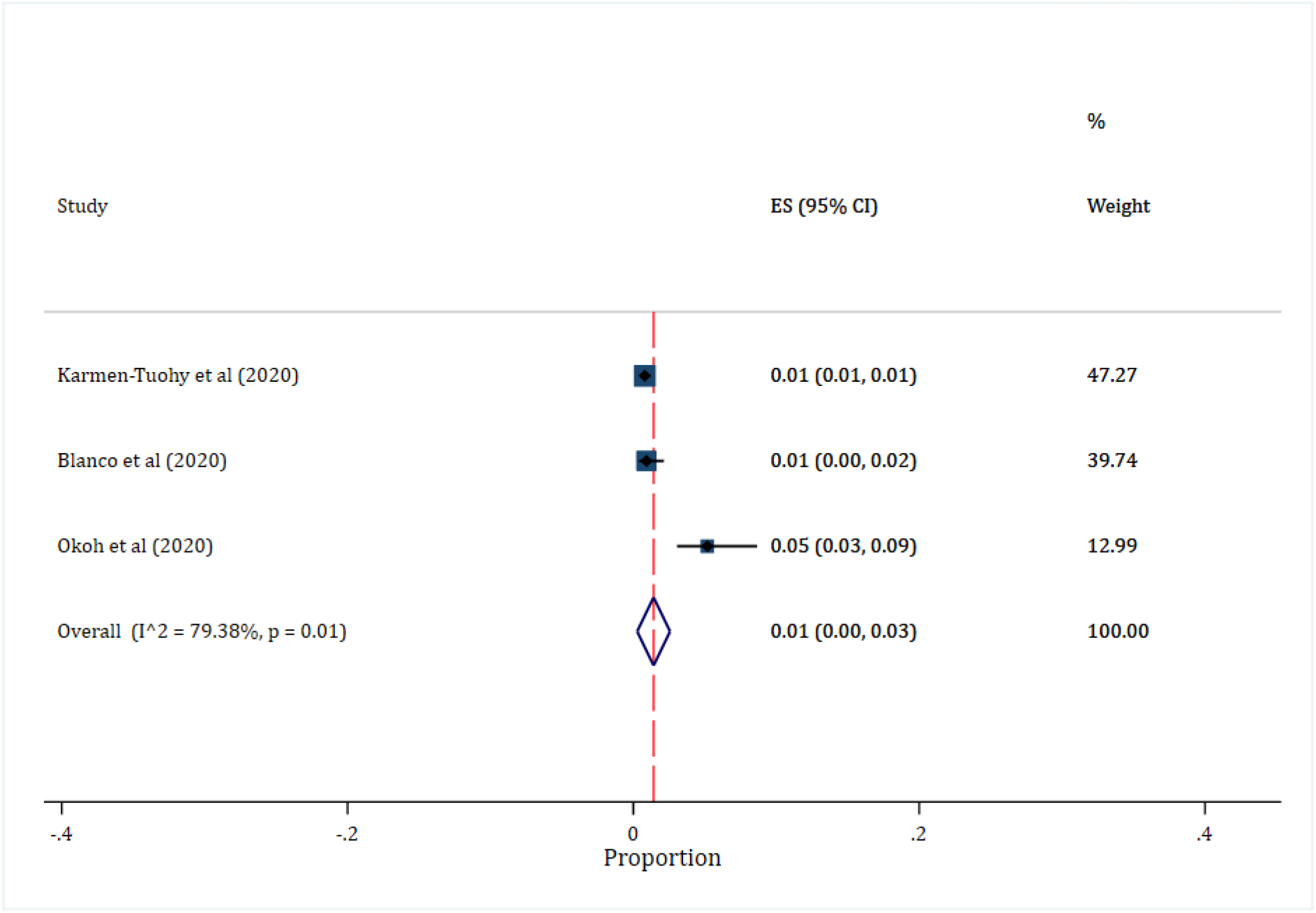
Pooled prevalence of HIV among SARS-CoV-2 cases

### Clinical features of HIV/SARS-CoV-2 co-infection

#### Demographic characteristics

Of 133 patients, 112 (84.2%) were male whereas 2 were transgender. One study did not report the sex of PLHIV (8). Five participants were men who have sex with men (9,18). The higher proportion of males with COVID-19 could partly be explained by lower activity of innate and adaptive immune responses against viruses compared to women (19). The mean/median age (range) was 30 – 60 years (7,10,11,13). It appears that the majority of PLHIV in these studies were < 65 years, which would otherwise increase the risk for poor outcomes (20). Paediatric HIV/SARS-C0V-2 co-infection was not reported, and therefore an area for further investigation.

### Symptoms and other comorbid conditions

The commonest symptoms were fever (73.8%, n = 76) and cough (57.3%, n = 59). Four studies (7,8,13,21) did not report symptoms for 43 patients. The duration (range) of symptoms before presentation was 1 – 14 days (n = 16) (9,12,14,18,22–25). Fever and cough have been consistently reported to be the commonest symptoms of COVID – 19 and our finding is in agreement with this observation (26). Sixty two (65.3%) patients had at least one other comorbid condition (9–11,18,22,23,27,28). This is expectedly higher than the 25.1% reported among HIV negative individuals in China (29). Among the counts of comorbid conditions, the commonest were hypertension, hyperlipidaemia and diabetes mellitus. Similarly, hypertension (21.1%) and diabetes (9.7%) are the most prevalent comorbid conditions among HIV negative patients with COVID – 19 (2). Hyperlipidaemia has not been a key comorbidity in previous reports on COVID –19. It should however be expected among PLHIV since their lipid levels are reportedly higher compared to the general population (30). Interestingly, low levels of low-density lipoproteins (LDL) are associated with COVID – 19 disease progression and mortality (31). It is unclear whether hyperlipidaemia has a protective role against severe disease and outcomes of HIV/SARS co-infection. On the other hand, the co-infection with Hepatitis B and/or C (9.7%) that we observed could increase COVID – 19 severity and mortality (32). A total of 129 (97.0%) patients were antiretroviral therapy (ART) experienced, and among those with a known HIV viral load, 113 (85.6%) were suppressed. One study did not report ART status (8). Given the high ART coverage in this population, our findings should be interpreted in the context of ART experienced, virologically suppressed PLHIV. Only 3 patients were explicitly reported as ART naïve (9,27,28). The clinical characteristics of HIV/SARS-CoV-2 co-infection among ART naïve PLHIV remain largely unknown. Nevertheless, antiretroviral therapy seems not to have profound effect on SARS-CoV-2, although it is protective against superimposed opportunistic infections (33).

### Laboratory abnormalities

The median/mean most recent CD4 count (range) was 298 – 670 cells/mm^3^ (n = 107) (7,10,11,13). Studies reported elevated fibrinogen levels (12,27), C-reactive protein (7,9,12,14,18), D-dimers (9), lactate dehydrogenase (7,9,12) and liver enzymes (14,15,27) among individual patients. Thrombopenia (9,12), lymphopenia (9,12,14,28), higher lymphocyte count than HIV negative persons (7) and normal procalcitonin levels (7,9,12,14,25,27) were also reported. The counts were too few do derive meaningful interpretations.

### Radiological imaging abnormalities

CT and X-ray abnormalities were observed among 46 (54.1%) patients (7,9,10,14,22,23,25). This could be an underestimate since 56.5% of these were by X-ray, which has a low sensitivity among patients with COVID – 19 (34). Similar to studies among HIV negative SARS-CoV-2 cases (35), bilateral lung involvement was the commonest pattern.

### Disease severity and drugs used in the management of HIV/SARS-CoV-2 co-infection

Antibiotics, hydroxychloroquine, and lopinavir/ritonavir (as COVID-19 therapy, not ART) were prescribed for 27 (29.3%), 20 (21.7%), and 16 (17.4%) patients respectively. Treatments were not reported for 54 patients (8,11,13). Although the optimal treatment for COVID – 19 in HIV/SARS-CoV-2 co-infection is unknown, the safety and efficacy of lopinavir/ritonavir, hydroxychloroquine and antibiotics are still not well established (36). Randomised controlled trials are needed to determine the optimal ART among PLHIV with SARS-CoV-2. Disease severity was reported by 7 studies (9–11,13–15,25), and majority had mild disease (56.0%). Further, 80 (58.0%) of the HIV/SARS-CoV-2 cases were hospitalised. In contrast, 85.3% of inpatients with COVID –19 had moderate or severe disease in China (37). One study (13) did not report whether the patients were hospitalised or not. Table 2 shows other clinical features of HIV/SARS-CoV-2 co-infection.

**Table 2:**
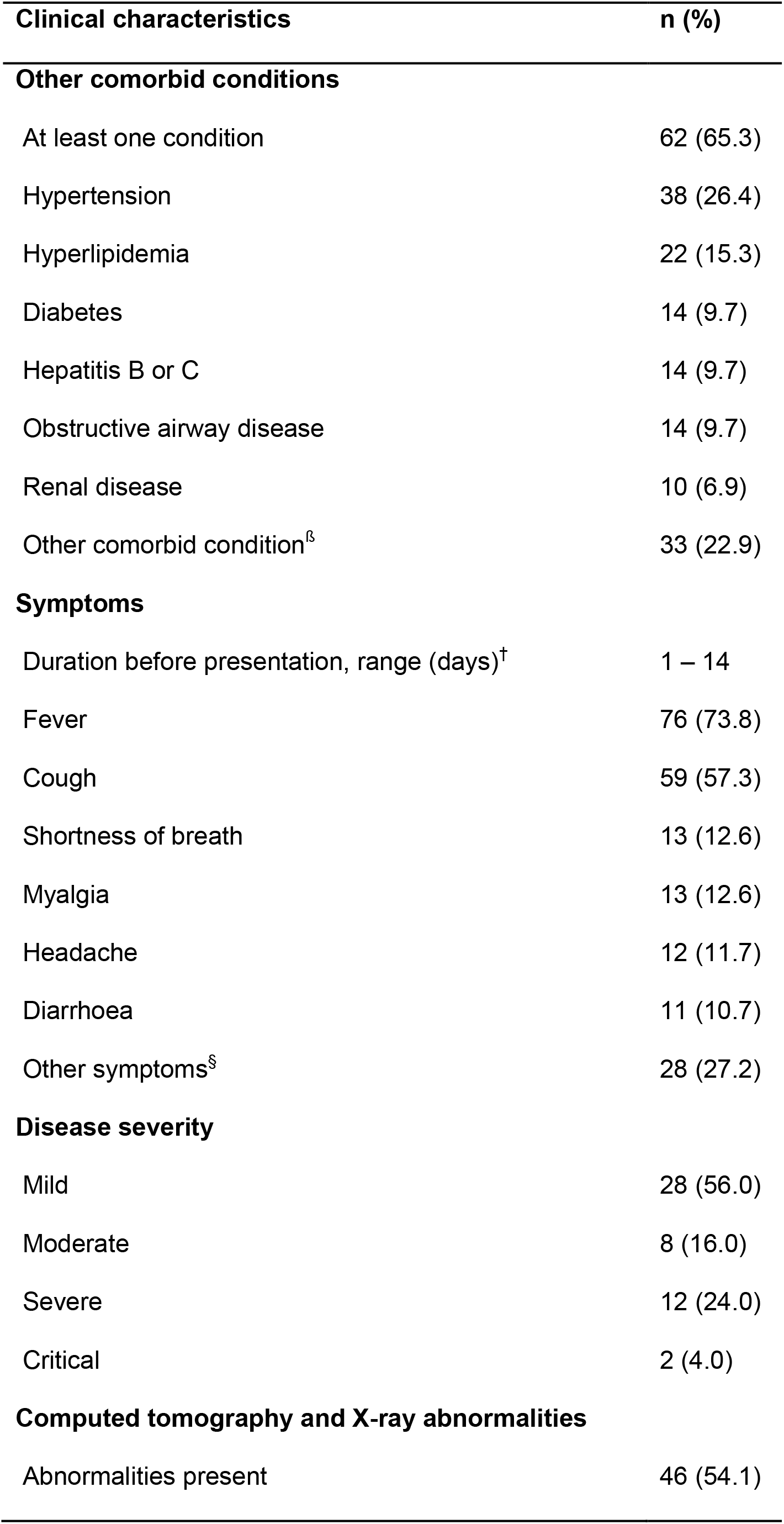
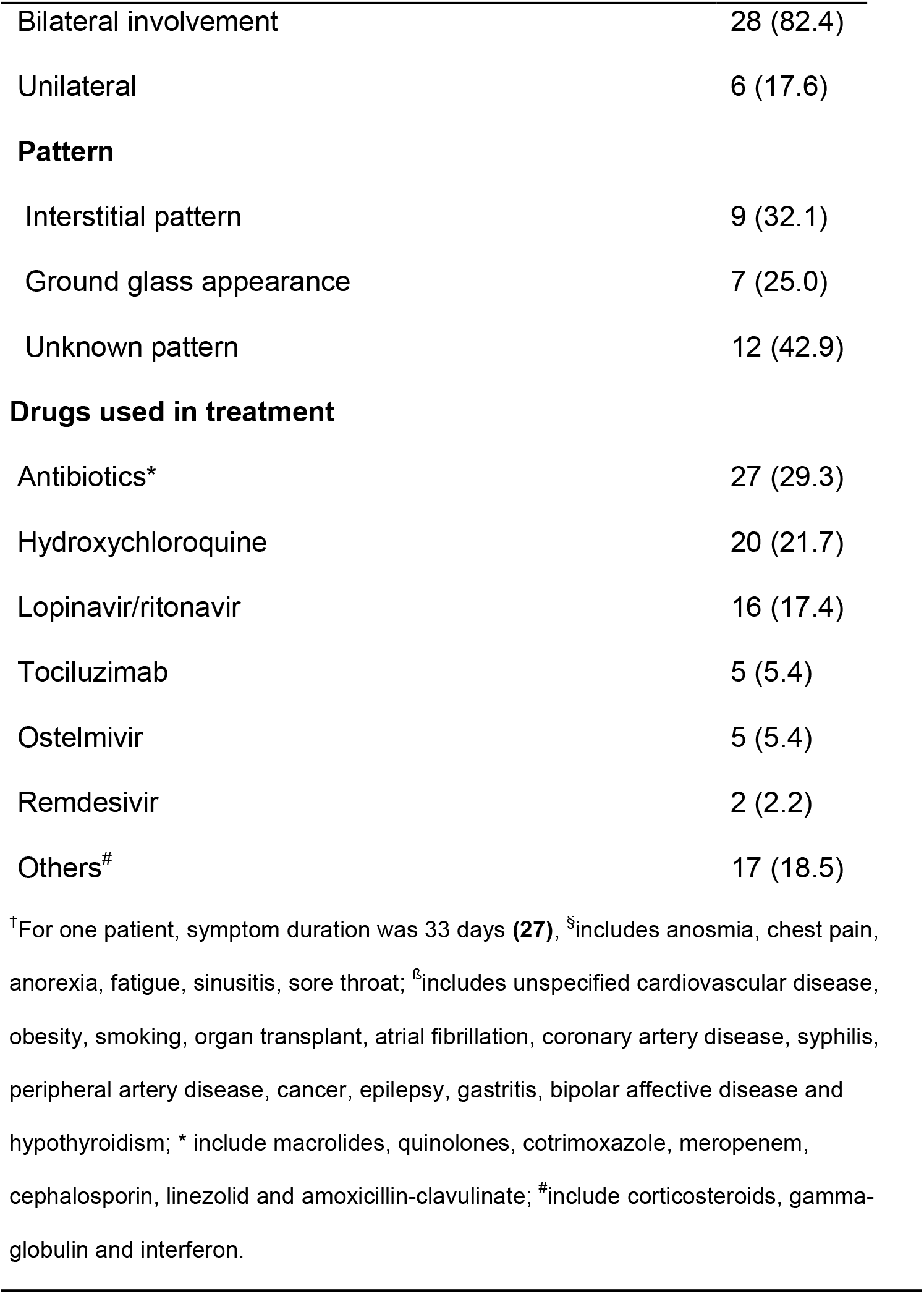
Clinical features of HIV and SARS-CoV-2 co-infection.

### Outcomes of HIV/SARS-CoV-2 co-infection

The length of hospital stay ranged from 1 – 24 days (7,9,15,18,25,27,28). Among patients with a known outcome, 120 (88.9%) recovered whereas 11.1% died. The pooled mortality, from 6 studies (7,8,10–13), was 9% (95% CI: 3.0 – 15.0, I^2^ = 25.6%, p = 0.24) (figure 3), well within the range of 3.5 – 13.9% reported elsewhere (38,39). The high recovery observed among HIV/SARS-CoV-2 co-infected patient could be explained by many factors. As described above, the population was mostly < 65 years of age with mostly mild disease. The possible protective role of hyperlipidaemia needs to be evaluated as well. Another hypothesis to explain the high recovery rate is that the impaired T-cell responses in HIV infection may be protective against severe lung inflammation (40). Okoh, *et al*., (8) also found HIV infection to have a survival benefit among minority populations with COVID – 19.

**Figure 3:**
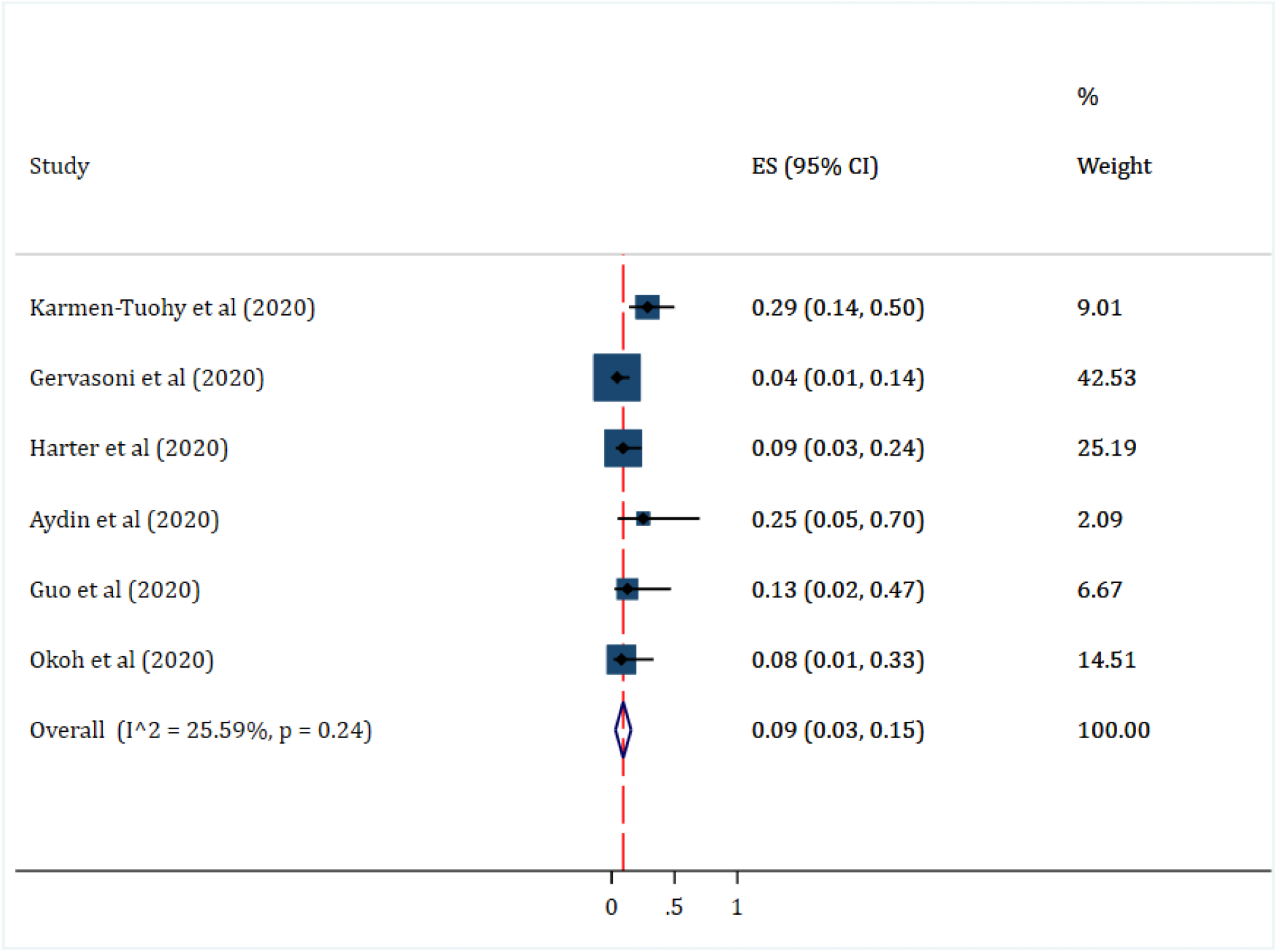
Pooled mortality among HIV/SARS-CoV-2 co-infection cases

There are key limitations of our study. First, the findings are not generalizable to low resource settings where the highest burden of HIV is found. Also, the majority of the studies reviewed were case reports and case series which inherently have bias. Additionally, the sample size is small and inferences cannot be made with certainty. Lastly, most of the patients in these studies were inpatients and therefore the reported severity of COVID –19 may not be accurate for the entire population of PLHIV. Larger studies are still needed to characterise the HIV/SARS-CoV-2 co-infection. Nevertheless, we have provided a preliminary but comprehensive synthesis of the current literature on HIV/SARS-CoV-2 co-infection.

## CONCLUSION

The prevalence of the HIV/SARS-CoV-2 co-infection is low. The clinical characteristics and outcomes of COVID – 19 among ART-experienced PLHIV and HIV negative individuals are comparable.

## Data Availability

Datasets used in this analysis are available from the corresponding author upon reasonable request

## TRANSPERANCY DECLARATIONS

### Conflict of interest

The authors do have any conflict of interest to declare.

### Source of funding

None

### Access to data

Datasets used in this analysis are available from the corresponding author upon reasonable request

## Acknowledgements

We appreciate all frontline healthcare workers taking care of COVID – 19 patients.

## Author contribution

JBB: Conception, design of the study, data acquisition, formal analysis, interpretation of data, drafting and revising article and final approval.
RO: Data acquisition, formal analysis, interpretation of data, revising article and final approval.
CA: Interpretation of data, revising article and final approval.
JN: Interpretation of data, revising article and final approval.
LR: Data acquisition, interpretation of data, revising article and final approval.
FB: Data acquisition, formal analysis, interpretation of data, revising article and final approval.
JN: Interpretation of data, revising article and final approval.

## APPENDIX 1 SEARCH STRATEGY

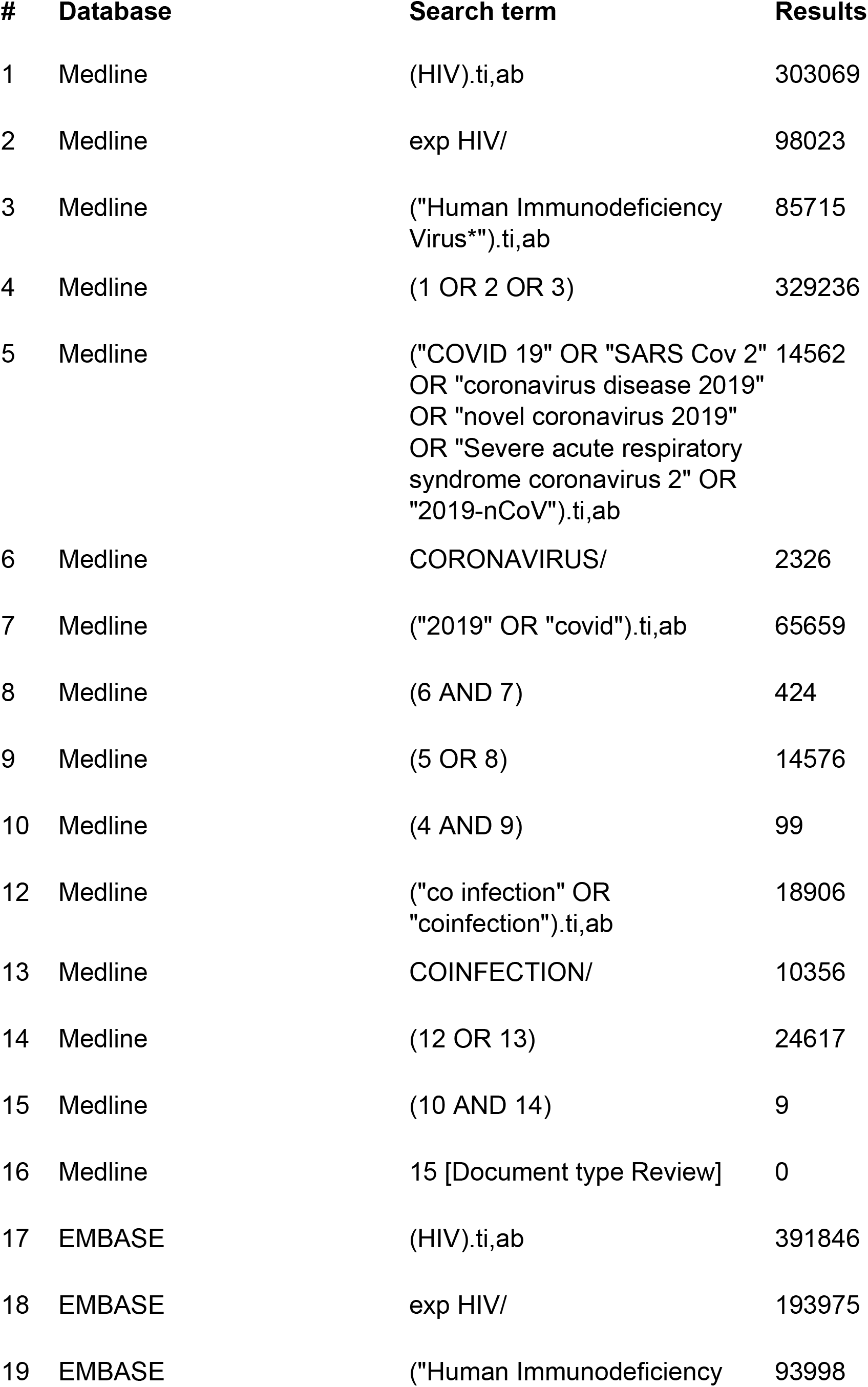
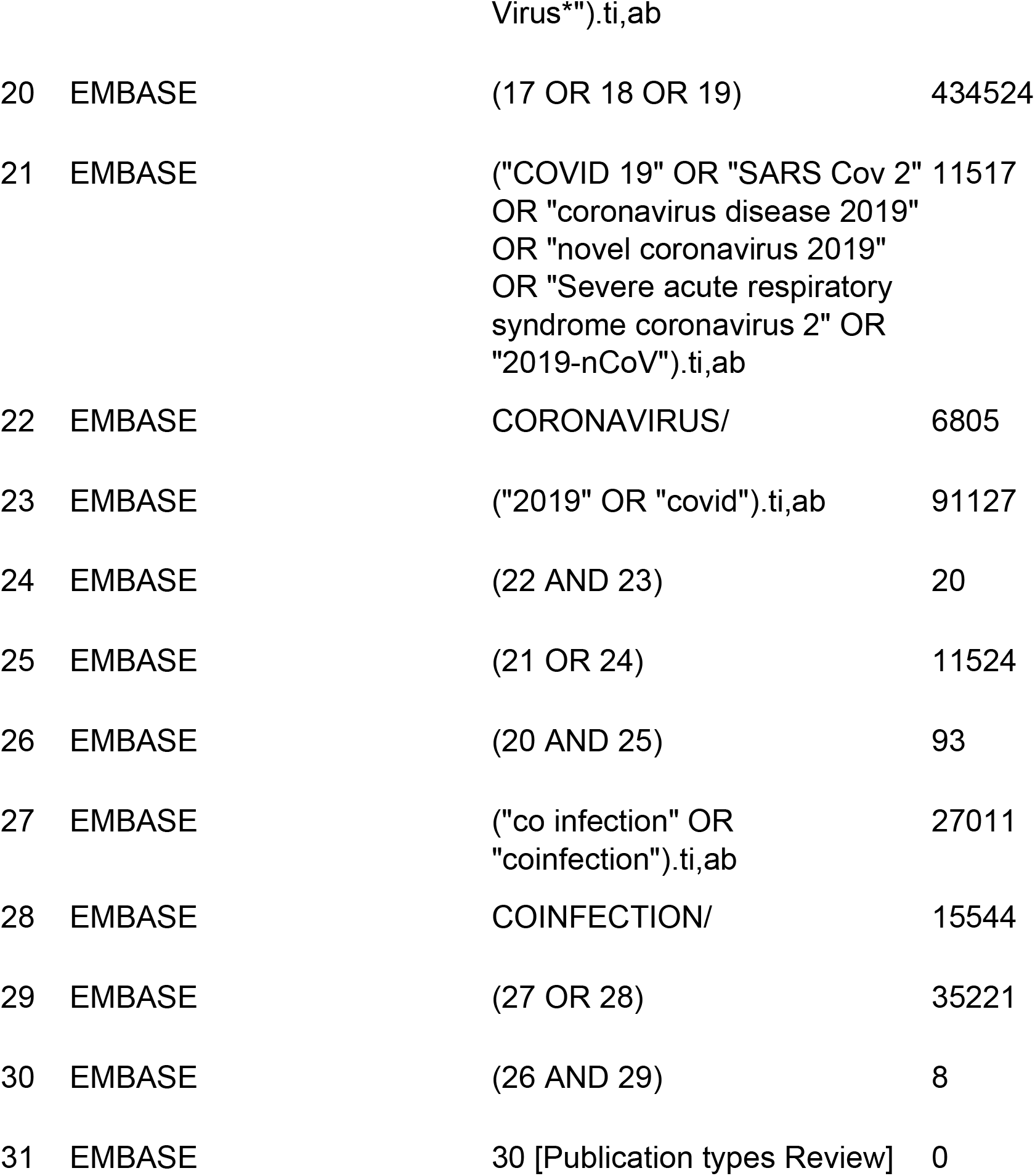

## Notes

### Competing Interest Statement

The authors have declared no competing interest.

### Funding Statement

There was no funding for this work.

### Author Declarations

The study is a systematic review and meta analysis of publicly available data. Ethical approval was therefore not sought

